# The onset of late severe lung impairment in COVID-19 is associated with high inflammation markers at admission and metabolic syndrome markers

**DOI:** 10.1101/2022.12.20.22282909

**Authors:** Olivier Epaulard, Audrey Le Gouellec, Marion Le Marechal, Benjamin Nemoz, Anne-Laure Borel, Anaïs Dartevel, Hubert Gheerbrant, Marie-Christine Herault, Annick Bosseray, Giovanna Clavarino, Julien Lupo, Damien Viglino, Fanny Quenard, Clara Candille, Boubou Camara, Michel Durand, Patrice Faure, Dorra Guergour, Elena Chidlovski, Marie-Christine Jacob, Sylvie Larrat, Marie Froidure, Nicolas Terzi, Sébastien Quetant, Jean-François Payen, Barbara Colombe, Tatiana Raskovalova, Patrice Morand, Isabelle Pierre, Carole Schwebel, Rebecca Hamidfar, Laurence Bouillet, Jean-Paul Brion, Candice Trocme, Sylvie Berthier, Carole Chirica, Anne-Laure Mounayar, Myriam Blanc, Patricia Pavese, Bertrand Toussaint

## Abstract

**Background:** COVID-19 severity is mainly related to lung impairment. However, preexisting patient characteristics and biomarkers at admission associated with this event are not precisely known.

**Methods:** We report 205 patients admitted for a proven COVID-19 in our institution between March 7 and April 22, 2020, particularly their comorbidities, respiratory severity, immune profile, and metabolic profile.

**Findings:** Median age was 70 years [interquartile range (IQR) 25-75: 60;79]; 115 (56·1%) patients were men. Oxygen supplementation of >2L/min was required in 107 patients (52·2%) after a median time of 8 days [IQR: 6;10] after the first symptoms; 67 (32·7%) patients were admitted to the intensive care unit (ICU), almost exclusively due to severe hypoxia. Patients requiring >2L/min oxygen therapy and/or ICU admission were older and more frequently males, with a significantly higher body mass index (BMI), a significantly higher total cholesterol (TC) / HDL cholesterol ratio, and higher triglycerides. They also had higher plasma levels of C-reactive protein (CRP) and interleukin 6 (IL-6); IL-6 >20 ng/L and CRP >70 mg/L were significantly associated with ICU admission and/or (for patients with a decision of limitation of life-support therapy) death. Higher BMI and TC/HDL-c ratio were associated with higher CRP and IL-6 levels. Steroid therapy was performed in 61 patients; while its clinical impact was inconclusive due to heterogeneous situations, IL-6 levels decreased significantly more in these patients.

**Interpretation:** Severe COVID-19 mostly relates to late-onset pneumonia associated with preexisting metabolic syndrome markers and a surge in inflammatory markers, allowing the early identification of at-risk patients.

**Funding:** This work was supported by Foundation University of Grenoble Alpes.

## Introduction

The pandemic of COVID-19 caused by the coronavirus SARS-CoV-2 has been responsible for 4,137,193 cases and 285,760 deaths worldwide as of May 12, 2020. The high case fatality observed in some areas (particularly with a high medical coverage such as Lombardy in Italy and Alsace in France) has been linked to the fact that a large proportion of patients with COVID-19 develop severe lung lesions and require admission to an intensive care unit (ICU) [1]. Indeed, previous studies reported that approximatively 25% of patients hospitalized with COVID-19-associated pneumonia required intensive care for respiratory support [2]. Due to the brutal epidemic profile, the number of ICU beds available in a given territory may soon be overwhelmed, leading to high mortality.

The lung impairment induced by SARS-CoV-2 is unique in several respects. First, it occurs at a frequency of 5% [1] to 10% [3], much greater than for other acute viral infections such as the influenza virus [4], respiratory syncytial virus (in adults) [5], and metapneumovirus [6] infections. Second, it has a late onset, as the median delay between the first symptoms and the need for ICU is 10 days [2]. Third, rare studies (to date) report that it is associated with a strikingly intense immune activation, sometimes referred to as “cytokine storm,” similar to the cytokine release syndrome that has been increasingly reported after the use of chimeric antigen receptor T cells [7]. In particular, higher interleukin 6 (IL-6) plasma levels were reported in patients with respiratory failure [8], along with high levels of inflammatory blood markers including C-reactive protein (CRP), ferritin, and D-dimers, an increased neutrophil-to-lymphocyte ratio, other cytokines such as IL-2R, IL-6, IL-8, IL-10, and tumor necrosis factor alpha (TNF-α) [9]. This raised the question as to whether immune-targeted therapy may be effective in preventing or shortening severe lung impairment.

The first cases of COVID-19 occurred in our area (Isere department, 1·3 million inhabitants) with a 2-week delay compared with other French regions. This allowed us to better assess the inflammatory phase, which began to be informally described at this time in patients admitted for hypoxemia due to SARS-CoV-2 infection.

In this study, we aim to report the clinical characteristics and immune profile of patients admitted in our institution for a proven SARS-CoV-2 infection of varying severity. Additionally, we aimed to assess the evolution of inflammatory markers in patients with or without steroid therapy.

## Methods

### Patient data

Our institution (Centre Hospitalier Universitaire Grenoble-Alpes) is a public university hospital with 2,100 beds in an area of 1·3 million inhabitants. Except for three patients hospitalized for nearly asymptomatic SARS-CoV-2 infection between February 8 and 21 (for isolation purposes) [10], the first patient who tested positive for SARS-CoV-2 in our area presented on March 7. This study includes all adult patients consecutively admitted for COVID-19 confirmed by reverse transcription polymerase chain reaction (RT-PCR) on respiratory samples (nasosinus swab, sputum, or tracheal aspirate) between March 7 and April 22, 2020. Patients admitted to the emergency department who did not require further hospital stay were not included. Patient demographics, clinical data, oxygen requirements, ICU admission, steroid treatment, and laboratory data from the hospital’s biochemistry, immunology and hematology departments, including serial samples, were extracted from electronic medical records.

### SARS-CoV-2 RT-PCR

RT-PCR was performed using several techniques due to shortages in reagent supplies. The first method implemented in our laboratory was developed by the French Pasteur Institute in Paris, which acts as the national reference laboratory for coronavirus infections. Two sets of primers targeting two regions in the RdRp gene allow for the detection of SARS-CoV-2. The second technique uses the R-gene® kit (BioMérieux, Marcy l’Etoile, France), which targets both RdRp and N genes. All extractions were performed using the eMag® system from BioMérieux. Amplification and detection were performed in the LightCycler 480 (Roche, Manheim, Germany) system.

### Interleukine measure

The analysis of IL-6 was performed on EDTA plasma samples using a magnetic bead-based multiplex immunoassay (High-sensitivity T cell panel reference, HSTCMAG-28SK) (Merck, Darmstadt, Germany) following the manufacturer’s instructions. Data from the reactions were acquired using the MAGPIX system (Luminex Corporation, Austin, TX, USA).

### Ethics

This study was conducted in accordance with the French legislation on the retrospective use of healthcare data, and ethical approval was given by the Clinical Research Direction of the Centre Hospitalier Universitaire Grenoble-Alpes. The BIOMARCOVID project is a research project that does not involve the human being and whose controller is the Centre Hospitalier Grenoble-Alpes. This research has been registered in accordance with French regulations and meets the requirements of the National Commission for Information Technology and Civil Liberties (CNIL: La Commission Nationale de l’Informatique et des Libertés) reference methodology 004.

### Statistical analysis

Mann-Whitney test was used to compare the distribution of values in two groups. Chi2 test was used to compare bimodal parameters in two groups. Log-rank test was used to compare the time to reach an event in two groups.

### Funding

This work was supported by the Foundation University of Grenoble Alpes.

## Results

### Population

Between March 7 and April 22 2020, 205 patients were hospitalized in our institution for a proven COVID-19 (positive RT-PCR) (figure 1). Patients were referred to COVID-19-dedicated units because of marked respiratory impairment, alteration of general status, or practical reasons (e.g., COVID-19 diagnosis in a psychiatry ward with patients subsequently transferred to ensure isolation procedures). Mean and median age were respectively 69·0±15 and 70 years [interquartile range (IQR) 25-75: 60;79]; 115 (56·1%) patients were men.

**Figure 1.**
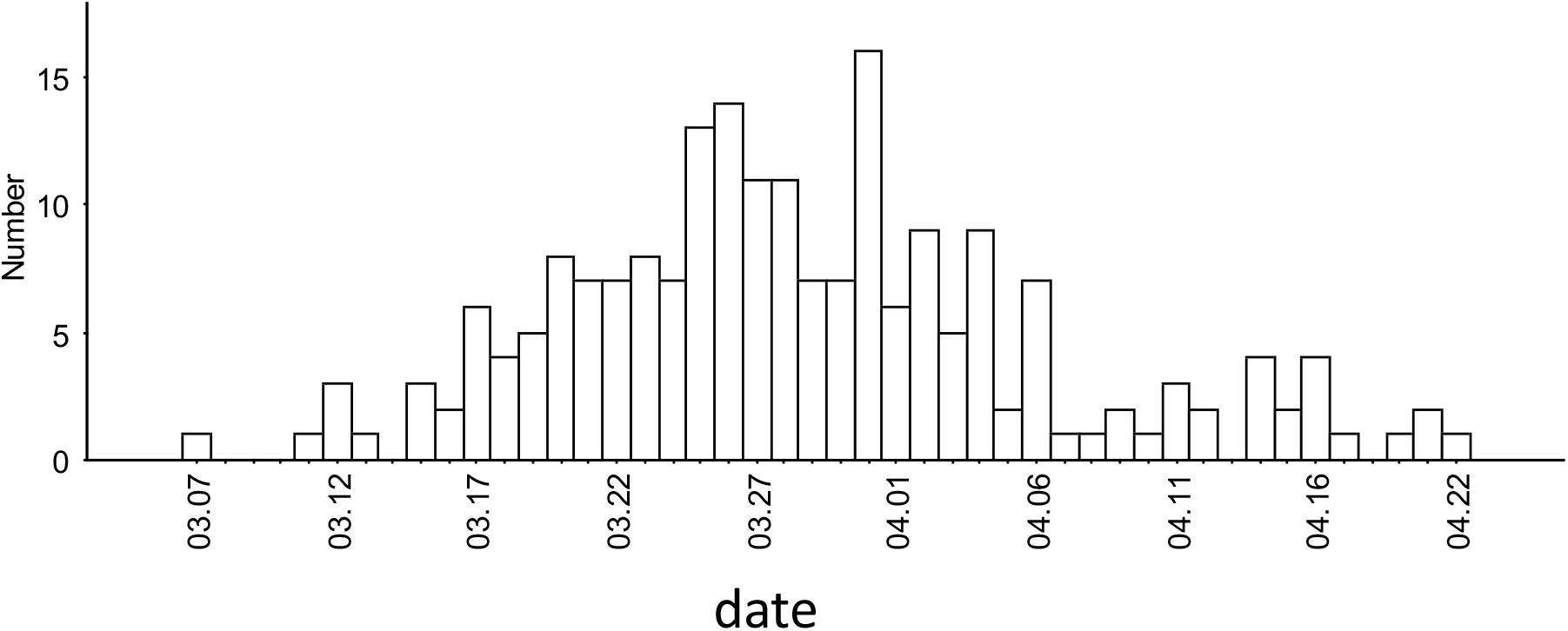
Admissions during the study period.

Oxygen supplementation was required in 150 patients (73·2%), including 107 (52·2%) >2L/min, after a median time of 8 days [IQR: 6;10] after the first symptoms and a median time of 0 days [IQR: 0;1] after hospital admission. A total of 67 (32·7%) patients were admitted to ICU after a median time of 8 days [IQR: 7;11] after the first symptoms and a median time of 1 day [IQR: 0;3] after hospital admission. All ICU admissions were due to severe acute respiratory deterioration except for one case of bacterial necrotizing fasciitis (unrelated to COVID-19) and one case of Guillain-Barre syndrome.

In two cases, the use of extracorporeal membrane oxygenation was required. Fifteen (7.3%) further patients with severe lung impairment were not referred to ICU due to a consensual decision for the limitation of life-support techniques (LLST) because of advanced age and severe comorbidities. Among the 205 patients, 22 (10·7%) died before April 30, including 6 in ICU and 15 patients with LLST; 16 other patients were still in ICU at the same date. Death occurred after a median of 10 days [IQR: 8;20] after hospital admission.

Table 1 presents the characteristics of patients with different severity classes (no oxygen required; oxygen required ≤2L/min; oxygen required >2L/min but no ICU admission; ICU admission; LLST). Patients requiring high (>2L/min) oxygen flow and/or ICU were older and were more frequently males. They also had a significantly higher body mass index (BMI), total cholesterol (TC)/high-density lipoprotein cholesterol (HDL-C) ratio, and higher triglyceride levels.

**Table 1.** Characteristics of patients in five groups according to COVID-19 severity

|  |  | Severity groups |  |  |  |  |  |
| --- | --- | --- | --- | --- | --- | --- | --- |
|  |  | No oxygen required<br>(N=55) | Oxygen required ≤2L/min<br>(N=41) | Oxygen required >2L/min but no ICU<br>required<br>(N=27) | Severe lung impairment requiring<br>ICU<br>(N=65) | Severe lung impairment but limitation<br>of life support therapy (N=15) | P (no oxygen required or ≤2L/min vs<br>other groups) |
| **Males (%)** |  | 20 (36.4%) | 24 (58.5%) | 9 (33%) | 52 (77.2%) | 10 (66.7%) | 0.005 |
| **Age\*** |  | 71 [54;83] | 67 [59;75] | 73 [67;83] | 67 [59;72] | 86 [76;90] | 0.546 |
| **BMI\***<br>(in 153 patients) |  | 24.1 [21;27] | 25.5 [24;28] | 31.0 [25;35] | 28.0 [25;30] | 27.9 [25;30] | <0.001 |
| **Diabetes (%) (when known)**<br>(in 126 patients) |  | 26.9% | 26.3% | 40.0% | 18.2% | 50.0% | 0.999 |
| **Length of hospital stay\*** |  | 6 [4;10] | 8 [6;10] | 9 [7;12] | 23 [16;33] | 8 [6;10] | - |
| **First CRP measure (mg/L)\***<br>(in 201 patients) |  | 36 [8;73] | 55 [28;83] | 63 [47;154] | 134 [83;204] | 122 [90;166] | <0.001 |
| **First IL-6 measure (ng/L) \***<br>(in 160 patients) |  | 7.5 [5;12] | 8.9 [6;15] | 13.8 [8;20] | 20.7 [8;67] | 18.6 [16;44] | <0.001 |
| **First IL-6 measure only if**<br>**performed in the first 96 hours**<br>**of hospitalization (ng/L)\***<br>(in 136 patients) |  | 7.7 [5;12] | 9.2 [6;17] | 13.8 [8;20] | 24.8 [13;70] | 18 [16;44] | <0.001 |
| **T Ly /mm<sup>3</sup>**<br>(in 76 patients) | **CD3** | 760 [628;1255] | 650 [8230;1130] | 630 [320;1040] | 445 [230;700] | - | <0.001 |
|  | **CD4** | 510 [362;787] | 470 [400;720] | 490 [210;540] | 290 [150;400] | - | <0.001 |
|  | **CD8** | 210 [185;388] | 180 [173;363] | 100 [60;200] | 130 [80;250] | - | <0.001 |
| **B Ly /mm<sup>3</sup>**<br>(in 76 patients) |  | 130 [93;175] | 80 [60;118] | 90 [30;160] | 100 [70;180] | - | 0.547 |
| **NK Ly /mm<sup>3</sup>**<br>(in 76 patients) |  | 120 [85;172] | 120 [60;268] | 110 [60;170] | 70 [40;140] | - | 0.031 |
| **Glycaemia at admission\***<br>(in 54 patients) |  | 4.8 [4.5;5.5] | 5.0 [4.7;6.0] | 6.4 [5.4;7.0] | 6.3 [5.7;7.2] | - | 0.12 |
| **Ratio (TC/HDL-C)\***<br>(in 73 patients) |  | 4.5 [3.5;4.9] | 4.5 [3.0;5.4] | 5.2 [5.0;6.2] | 5.5 [4.6;6.2] | - | 0.002 |
| **Triglycerides (g/L) \***<br>(in 92 patients) |  | 1.3 [1.1;1.6] | 1.5 [1.0;1.8] | 1.8 [1.3;2.1] | 1.6 [1.3;2.2] | - | 0.024 |

\*median and IQR. BMI, body mass index; ICU, intensive care unit; Ly, lymphocytes TC, total cholesterol, HDL-C: HDL-cholesterol.
The 2 patients who were admitted in ICU for a non-respiratory issue were not included in this table

### Immune profiles

Inflammation and immune system alterations were observed, with their intensity increasing with the severity of lung impairment across the five severity classes (table 1). In particular, CRP at admission and IL-6 at first measure (figure 2) were higher in more severe patients. Patients with higher initial plasma levels of CRP and IL-6 (particularly for patients who had this measure taken in the first 4 days of their hospital stay) required oxygen therapy more frequently. IL-6 >20 ng/L and CRP >70 mg/L were significantly associated with ICU admission and/or (for patients with LLST) death (figures 3). The follow-up of CRP and IL-6 showed that the intensity of inflammatory syndrome peaked at day 10 after the first symptoms; patients requiring high (>2L/min) oxygen flow and/or ICU reached higher values than the others (figures 4a and 4b)

**Figure 2:**
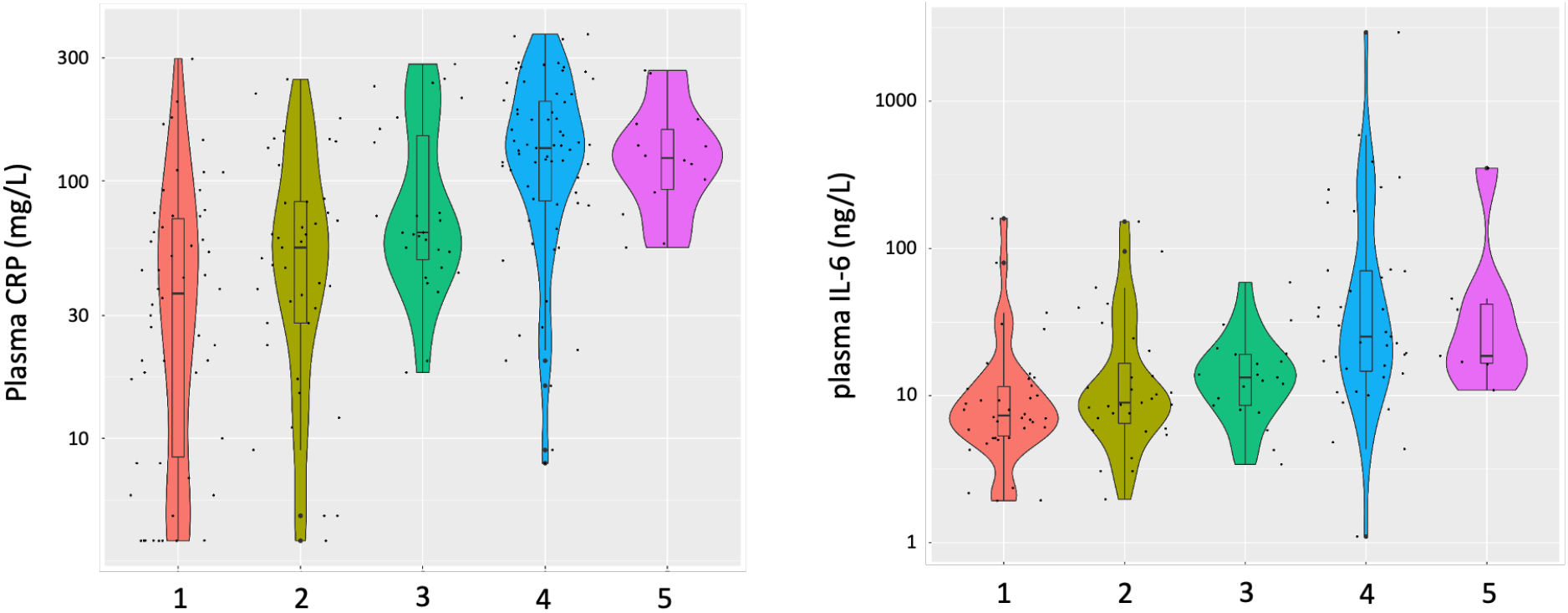
Distribution of the CRP at admission and of the initial IL-6 measure according to the five groups of severity (1 : no oxygen required, 2: oxygen :≤2L/min, 3: oxygen >2L/min but no ICU, 4: ICU, 5: LLST)

**Figure 3:**
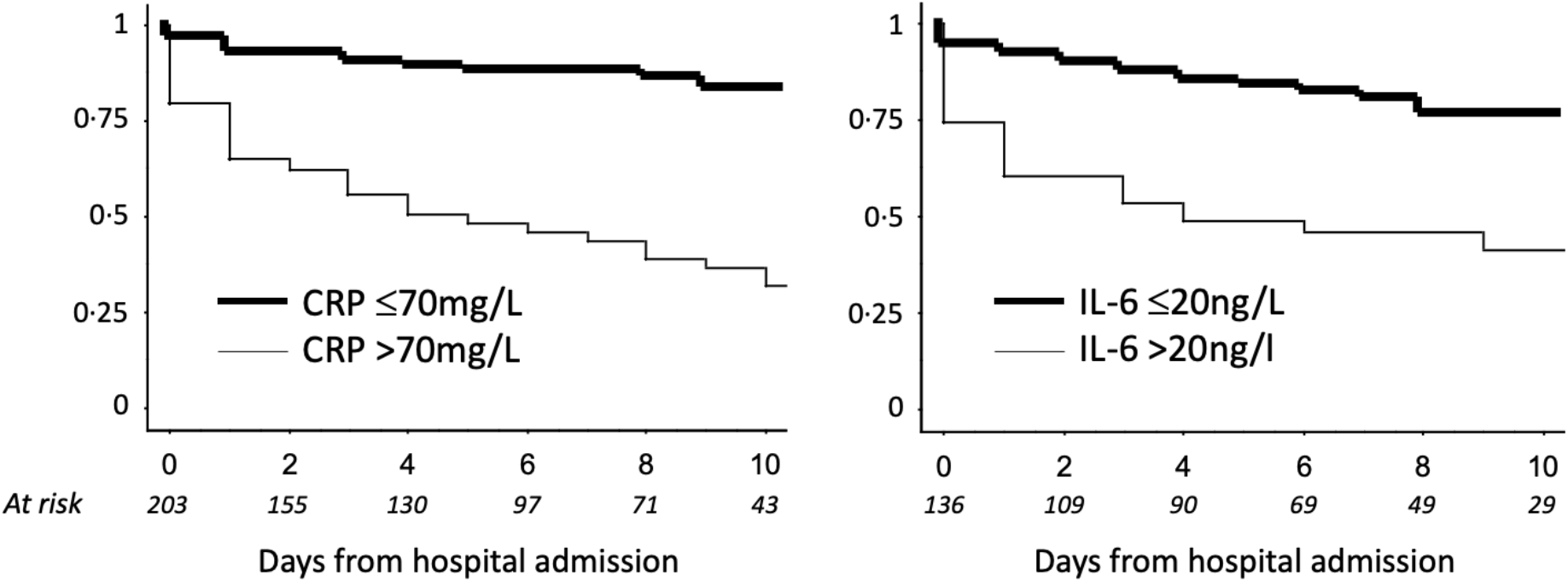
Time to ICU admission and/or to death (after LLST decision) according to the CRP at admission and the initial IL-6 measure (if performed in the 4 days from admission) (p<0.001 for both markers).

**Figure 4:**
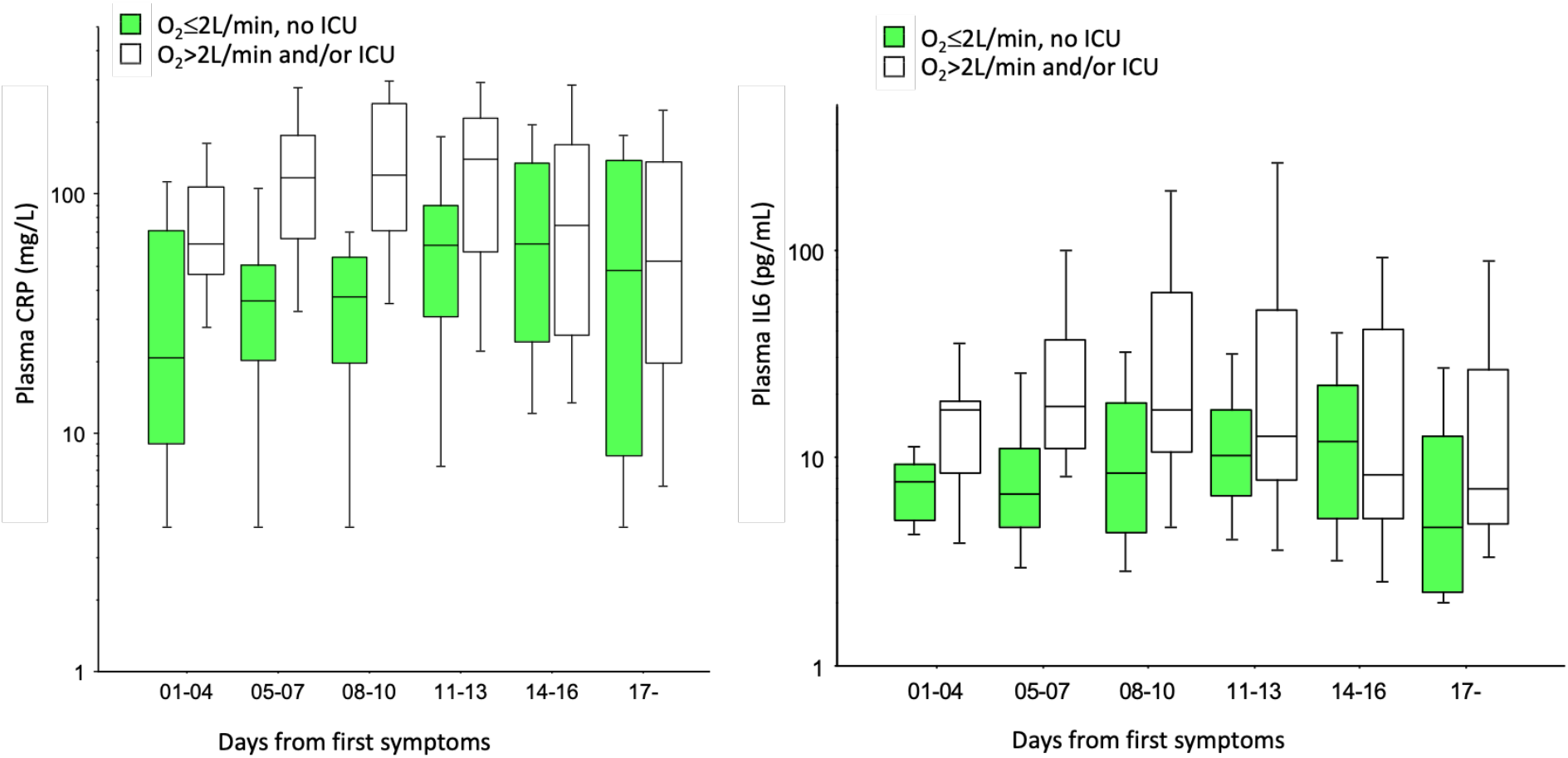
CRP measures (4a) and IL-6 measures (4b) over time according to severity (median and IQR in two severity groups)

Moreover, patients who required high oxygen flow and/or ICU had a more severe T lymphopenia (CD3+, CD4+ and CD4+ lymphocytes) and NK lymphopenia (table 1). They also had higher D-dimers and ferritin plasma levels (suppl. figures).

Of note, the median IL-6 plasma level was lower in women than in men (7·7 [5;17] vs 16.3 [10;40], p<0.001); it was also lower in patients with BMI ≤25 kg/m^2^ (8·9 [5;17] vs 14·0 [8;40], p=0·0134) and in patients with TC/HDL-C ratio ≤4 (7·8 [5;13] vs 16·1 [8;40], p=0.0235). In parallel, the median CRP plasma level was lower in patients with TC/HDL-C ratio ≤4 (35 [16;75] vs 110 [52;153], p=0.003).

### Therapy

At the time when we began to admit COVID-19 patients to our institution (March 7), very few scientific publications mentioned that late-onset (7 days after first symptoms) severe lung impairment due to COVID-19 was associated with alterations in the inflammatory/immune profile. Following the online publication of position papers in March 2020 in which clinical immunologists discussed the potential interest of immunomodulating therapy at this phase [11, 12], we chose to discuss the use of steroids in our institution to lower the inflammation/immune activation for each patient who required oxygen therapy and had CRP >50mg/l and/or IL-6 >20 ng/L. Finally, 61 patients (29.7%) received steroids (IV dexamethasone 20 mg/day for 5 days followed by 10 mg/day for 5 days, or oral prednisone or IV methylprednisolone 1 mg/kg/day for 5 days followed by 0·5 mg/kg/day for 5 days). Steroids were introduced at median time of 5 days [2;7] after hospital admission, including 50 ICU patients at a median time of 3 days [0;6] after ICU admission.

Table 2 shows the different situations regarding steroid therapy and outcome. However, it was not possible to establish the clinical impact of steroids, because of the heterogeneous nature of the situations.

**Table 2.** Outcomes of patients according to steroid therapy

|  |  | Situation at day 21 after admission |  |  |
| --- | --- | --- | --- | --- |
|  |  | Deceased | Still in acute care | Out of acute care |
| **No ICU; no LLTS** | **Steroids (N=8)** | 0 | 0 | 8 |
|  | **No steroids (N=110)** | 1 | 0 | 109 |
| **LLST** | **Steroids (N=3)** | 3 | 0 | 0 |
|  | **No steroids (N=12)** | 12 | 0 | 0 |
| **Later admission to ICU** | **Steroids before ICU admission (N=2)** | 0 | 1 | 1 |
|  | **Steroids between ICU admission and day 2 of ICU (N=6)** | 0 | 2 | 4 |
|  | **Steroids after day 2 of ICU (N=5)** | 1 | 1 | 3 |
|  | **No steroids (N=4)** | 0 | 0 | 4 |
| **Direct admission to ICU** | **Steroids between ICU admission and day 2 of ICU (N=12)** | 0 | 3 | 9 |
|  | **Steroids after day 2 of ICU (N=25)** | 0 | 10 | 15 |
|  | **No steroids (N=10)** | 1 | 2 | 7 |

*We excluded two patients who were admitted to ICU for reasons other than lung impairment as well as six patients who received various doses of steroids for different conditions before the COVID-19 diagnosis or who received a very short heterogeneous course of steroids during the hospital stay due to a supposed allergy. ICU, intensive care unit. LLST, limitation of life support therapy.*

Among the 26 patients who received steroids between the two measures of plasma IL-6, a median decrease in the level of this cytokine of 42·5 [10;126] was observed, compared with a median decrease of 0·2 [-3;6] in 82 patients who did not receive steroids between the two IL-6 measures (figure 5) (in four other patients, steroids were introduced before the first IL-6 measure).

**Figure 5:**
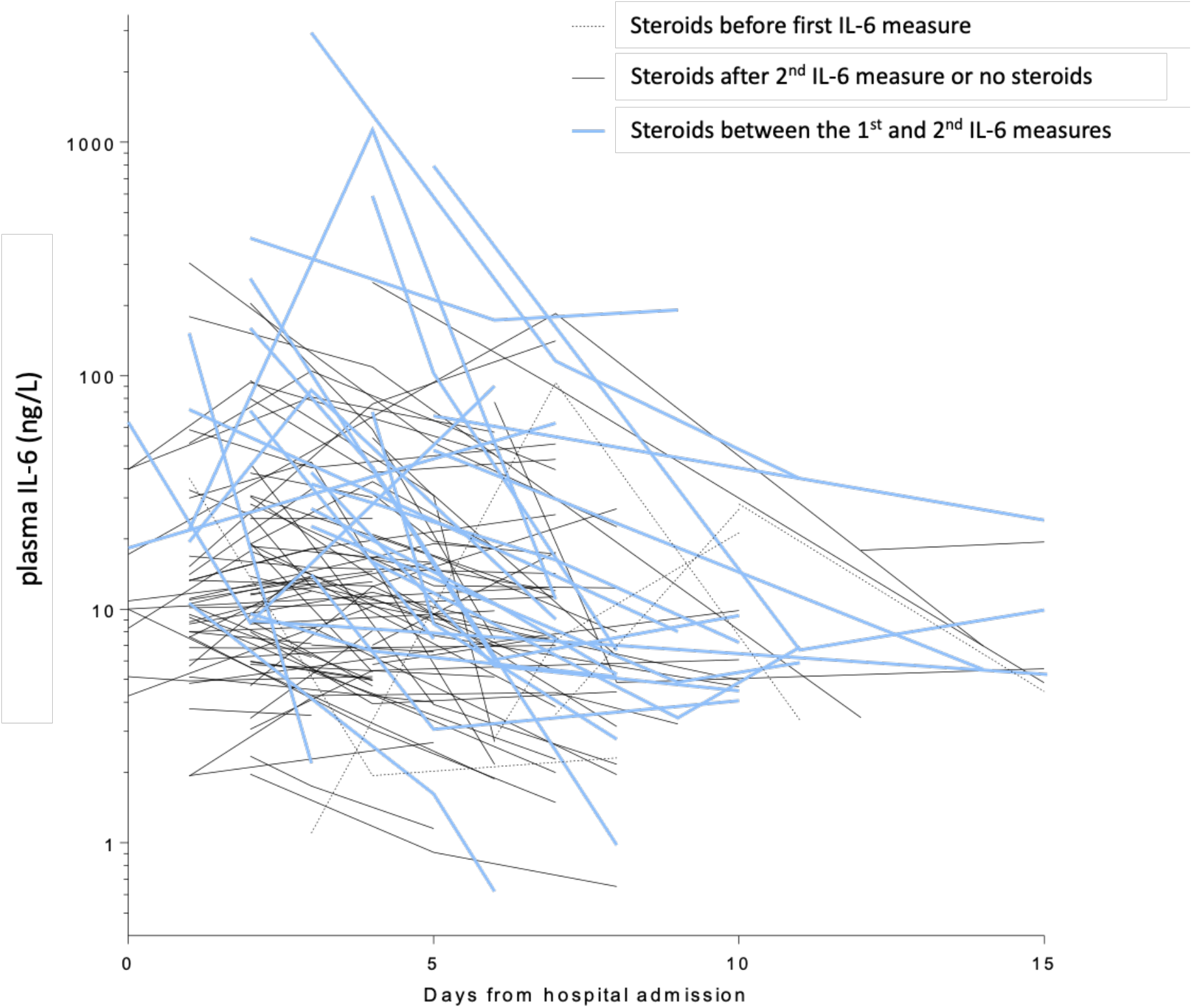
IL-6 kinetics under steroids for the 111 patients with at least 2 measures (one line per patient)

## Discussion

SARS-CoV-2 is the third documented zoonotic coronavirus that established an infection in a large number of human beings after SARS in 2002-2003 (8,096 cases) [13] and MERS since 2012 (2,494 cumulative cases in December 2019 (https://www.who.int/emergencies/mers-cov/en/, accessed April 30 2020); it is also the most damaging in terms of the number of infected people and mortality. The high case fatality rate might not be unexpected considering the figures in SARS and MERS (approximately 10% and 34%, respectively). However, the fact that the fatality rate is at least 10 times higher than that of other respiratory viral infections (excluding avian influenza) may partly explain why the initial strategies of most European and Northern American countries have sometimes been described as insufficient [14].

As previously reported in specific situations such as the Diamond Princess cruise ship cluster [15] (which allowed the detection of all infected subjects, even those with no or only benign symptoms), we observed heterogeneous clinical presentations, from nearly asymptomatic infection to extreme lung impairment requiring extracorporeal membrane oxygenation. Among others, male sex and higher BMI were associated with a more severe infection. High BMI is a well-known risk factor for severe influenza [4], although the impact of sex is less clear in the case of influenza [16]. In the largest (72,314) case series of COVID-19 to date [17], mortality was higher in males than in females (2·8% vs 1·7%); moreover, in a report of 1,591 patients admitted to ICU for COVID-19 in Italy, 82% were males [3]. However, the rate of symptomatic infection did not differ according to sex in the Diamond Princess cruise ship study. This link with sex may be related to the fact that men are more likely to have chronic lung or cardiovascular diseases, and therefore a higher risk of poor tolerance to infection. Interestingly, BMI did not differ between males and females in our study (data not shown).

We observed an association of markers of metabolic syndrome (high BMI, high TC/HDL-c ratio, high triglyceride levels) with COVID-19 induced inflammation, and with COVID-19 outcomes BMI is a known risk factor not only for severe cases of pandemic 2009 H1N1 influenza [18], but also for severe MERS cases [19]. This may be related to the restrictive pattern and reduced lung volumes in obese individuals; however, it may also be related to their fat distribution. Indeed, visceral fat, as opposed to subcutaneous fat, produces IL-6 and TNF-α, leading to low-grade inflammation [20]. The visceral fat accumulation that characterizes abdominal obesity is more frequent in men and is associated with comorbidities such as type 2 diabetes, hypertension, and non-alcoholic steatohepatitis. Some of these characteristics (male sex, excess body weight) have been recently identified as predictive factors of more severe COVID-19 cases. In a Chinese study, BMI >25 kg/m^2^ was associated with a six-fold increased risk of severe COVID-19 [21]; in a French study, 47·6% and 28·2% of cases requiring mechanical ventilation had a BMI >30 kg/m^2^ and >35 kg/m^2^, respectively [22]. These works and the present suggest that patients with a high BMI and elevated triglycerides and TC/HDL-C ratio (which characterize the lipid profile of metabolic syndrome) should be closely monitored in case of SARS-CoV-2 infection, and should be a key population for vaccine campaigns if a vaccine becomes available, and for early antiviral treatment if such a therapy is proven efficient.

We observed that patients with COVID-19 presented with complications that occurred late in the infection timeline, after a median delay of 8 days after the first symptoms; moreover, increased severity was associated with increased alterations of their inflammatory/immune profile, with higher plasma IL-6 and CRP and lower blood T lymphocyte count. This particular pattern has been gradually acknowledged in COVID-19 [8]. Interestingly, this was already reported in a study on 2003 SRAS patients [23] with an increase in IFN-γ, CXCL9/MIG, and CXCL10/IP-10 during the first 4 days of fever, followed by IL-8, IL-18, IL-6, and TGF-β; MIG, IP-10, and IL-18 levels were correlated with outcome. The term “cytokine storm” had already been coined in 2017 to describe the mechanisms of severe SARS and MERS [24]. It was proposed that this excessive deleterious immune response results from an incompletely controlled viral replication and delayed interferon responses, leading to the onset of an inappropriate inflammation at the primary site of the viral infection (i.e., the lungs), although other inflammation sites (including myocarditis [25] and nephritis [26]) have been reported during the COVID-19 epidemic. Our study suggests that IL-6 and CRP may be used to identify patients at higher risk of lung impairment who require closer respiratory monitoring.

In accordance with these reports, it was proposed that immunomodulatory therapy should be discussed for patients who require oxygen therapy and present such an immune activation. Caution is nevertheless required, as steroid therapy in MERS was associated with poorer outcome [27] and was inconsistently associated with positive results in 2003 SARS patients [28]. Warnings were first issued regarding the deleterious effect of dexamethasone on COVID-19 ARDS [29]. Conversely, immune intervention was successfully reported with a monoclonal antibody directed against the IL-6 receptor tocilizumab [30]. Our results suggest that an IL-6-based intervention may have a positive effect. In the patients reported in this study, steroids were associated with a decrease in IL-6 plasma levels; due to the central role that this cytokine is likely to play in COVID-19-associated lung lesions, this decrease may be beneficial for patients. However, due to the heterogeneity of situations, it was not possible to draw conclusions regarding steroid impact on survival.

Our study has several limitations, the main one being that steroid therapy was not homogeneously applied due to the ongoing progress made in COVID-19 knowledge during the 6 weeks of the study. Patients with marked respiratory symptoms and inflammatory profiles did not systematically receive steroids in the early period or only several days after ICU admission. This prevents us from drawing firm conclusions regarding the prognostic impact of this therapy for COVID-19. Another limitation of this study is the absence of evaluation of potential antiviral therapies. Finally, the present work was only performed in our hospital, resulting in a retrospective observational monocentric study and thus moderating its conclusions.

## Conclusions

A notable proportion of individuals admitted for a SARS-CoV-2 infection develop late-onset severe lung impairment. In our study, these patients were more frequently males, with markers of metabolic syndrome, and higher inflammation markers. These findings may allow the early identification of patients who require careful in-hospital observation and lead to a more rational management of care resources. Such patients might also benefit from immunomodulatory therapy, although randomized trials are needed in this respect.

## Data Availability

All data produced in the present study are available upon reasonable request to the authors

## Acknowledgements

We thank Pierre Audoin and Ilhem Elamrani for their support regarding the reglementary procedures.

